# Phenome-wide HLA association landscape of 235,000 Finnish biobank participants

**DOI:** 10.1101/2020.10.26.20219899

**Authors:** Jarmo Ritari, Satu Koskela, Kati Hyvärinen, FinnGen, Jukka Partanen

## Abstract

The human leukocyte antigen (HLA) system is the single most important genetic susceptibility factor for many autoimmune diseases and immunological traits. However, in a range of clinical phenotypes the impact of HLA alleles or their combinations on the disease risk are not comprehensively understood.

For systematic population-level analysis of HLA-phenotype associations we imputed the alleles of classical HLA genes in a discovery cohort of 146,630 and replication cohort of 89,340 Finns of whom SNP genotype data and 3,355 disease phenotypes were available as part of the FinnGen project.

The results suggest HLA associations in phenotypes not reported previously and highlight interactions between HLA genes and alleles in autoimmune diseases. Furthermore, shared HLA alleles in autoimmune and infectious diseases support a genetic link between these diseases.

## Introduction

Regulation of adaptive immune system function is based on recognition of foreign antigens and infectious agents by human leukocyte antigen (HLA) receptors encoded by highly polymorphic loci within the major histocompatibility complex (MHC) on chromosome 6. Out of more than 200 genes harbored by the MHC region approximately half have known immune-related functions (The MHC sequencing consortium, 1999). The HLA molecules play a key role in the initiation of immune response by binding internal (HLA class I molecules *A, B, C*) and external (HLA class II molecules *DR, DQ, DP*) peptides and presenting them to T lymphocytes. While class I receptors present antigens directly to cytotoxic CD8+ T cells, the class II molecules are recognized by CD4+ T cells that polarize into different regulatory subtypes (A. Barr et al., 2012). The extremely high genetic polymorphism of HLA genes results in structural variation in the peptide binding pockets between HLA alleles, consequently leading to different peptide-binding preferences and varying antigen repertoires presented to T cells.

Originally discovered over 50 years ago as the major determinant of organ and hematopoietic graft rejection (Thorsby, 2009), genetic variation in HLA has since been linked to a wide spectrum of immunological diseases (Trowsdale and Knight, 2013). In major multifactorial autoimmune diseases, HLA alleles and their protein-level motifs present the most important single genetic component in disease susceptibility (Matzaraki et al., 2017), even though in most diseases the triggering peptide complexing with the implicated HLA protein polymorphism remains unknown (Dendrou et al., 2018). On the other hand, varying degrees of protective allelic effects as distinguished from the absence of strong susceptibility alleles have been reported for major autoimmune disorders (Bettencourt et al., 2015; van der Helm-van Mil et al., 2005; van Lummel et al., 2019). The effect towards the reduction of disease risk is presumably mediated through presentation a favourable selection of antigens in terms of specificity and self-regulation (Tsai and Santamaria, 2013). Accordingly, both susceptibility and resistance effects have been attributed to amino acid residues and their positions in the HLA protein sequence (Furukawa et al., 2017; Gregersen et al., 1987; Raychaudhuri et al., 2012). Different alleles sharing a similar structural motif also manifests in local epistasis. Detailed analyses of large cohorts of patients with rheumatoid arthritis or type 1 diabetes have demonstrated that the MHC-mediated risk can be pinpointed to specific amino acid positions, and the effect is being modified non-additively by amino acid polymorphisms in a few other positions in the same or different class II gene (Hu et al., 2015; Lenz et al., 2015; Okada et al., 2016).

HLA allelic variance can cause differences in the strength of immune response against infectious agents such as HIV by differential preference of viral peptides (The International HIV Controllers Study, 2010). However, in case of structural similarity between pathogen T cell epitope and a host peptide, immune reaction against the antigen may also increase the likelihood of developing autoimmunity (Oldstone, 1998). Predisposition to infections before the onset of an autoimmune condition has been reported in several cases (Sfriso et al., 2010), and reaction of host T cell clones against the pathogen epitope mimicking host structures has been demonstrated experimentally (Wucherpfennig and Strominger, 1995). Nevertheless, exposure to a rich microbial environment also contributes to achieving protective, tolerogenic setting through toll-like receptor, regulatory T cell and interleukin signaling (Bach, 2018). Immunological regulation and its perturbation are therefore dependent on both environmental and host genetic factors that are mediated by individually varying HLA presentation.

Large biobank genome data collections combined with electronic health records have made phenome-wide association studies (PheWAS) feasible (Denny et al., 2013), leading to increased power and novel discoveries in disease genetics (Diogo et al., 2018; Liu et al., 2016; Verma et al., 2018). Population-based approach for the analysis of phenotypic spectrum of HLA associations can give novel insights into the architecture of well-established autoimmune and immune disease associations and broaden the view toward other traits as well (Hirata et al., 2019; Karnes et al., 2017; Liu et al., 2016). The first reported HLA PheWAS analysis with over 11,000 individuals found eight novel phenotypes linked with MHC SNPs as well as five previously unknown associations across multiple phenotypes (Liu et al., 2016). Karnes and coworkers (2017) imputed HLA alleles from cohorts of 28,839 and 8,431 individuals of European origin and tested HLA associations with 1,368 phenotypes. 104 significant associations were observed with 29 phenotypes and 29 HLA alleles. In addition to well-established HLA associations, four novel phenotypes were reported. Hirata and coworkers (2019) analyzed 106 clinical phenotypes for association with MHC variation in a cohort of 166,190 individuals from Japan. They reported significant genotype–phenotype associations in 52 phenotypes, and their fine-mapping showed multiple different patterns of HLA associations, some of which were independent from classical HLA genes.

Here we report a systematic, population-based association study of imputed HLA alleles in 3,355 phenotypes in discovery and replication cohorts of 146,630 and 89,340 individuals, respectively. These large single-population cohorts enabled us to perform HLA analysis in diseases not studied in detail before and to reveal cross-phenotype dependencies of allelic associations particularly between autoimmune and infectious diseases. Furthermore, as a systematic examination of risk-modifying effects have not, to our knowledge, been implemented at biobank-scale to date, we sought to define protective allelic effects as opposed to nonpredisposition to the top risk alleles. To this end, we studied heterozygous risk allele genotypes, and hypothesized that a risk allele effect could also be modified by a HLA locus of a different class.

## Results

### Associations of imputed HLA alleles

Altogether 155 four-digit HLA alleles were imputed with posterior probability > 0.5, and of these, 84 alleles had at least one confirmed association in both cohorts. In total, we found 3,649 statistically significant HLA-allele-phenotype associations in 368 phenotypes (Supplementary Table 1). Supplementary Figure 1 summarises the distribution of allele associations across the main phenotype categories for each HLA gene. HLA class II genes harboured both the largest number of associations and the strongest associations as indicated by their effect sizes. The top disease categories in terms of number of associations were type 1 diabetes and rheumatic diseases. We did not find a relationship between the number of significant associations and the number of available cases in a phenotype (Supplementary Figure 2).

1,620 of the 3,649 replicated HLA associations were in diabetes-related traits (Supplementary Table 1) with *DQB1*\**03:02* as the top risk allele. Celiac disease (CD) had the second highest number of HLA associations. The lowest p-values were for *DRB1*\**03:01, DQA1*\**05:01* and *DQB1*\**02:01* followed by other alleles known to be in a strong linkage disequilibrium with this HLA class II haplotype.

To validate our analysis we compared our results with previously published HLA PheWAS studies (Hirata et al., 2019; Karnes et al., 2017; Liu et al., 2016). We observed a consistent relationship between the obtained odds ratios of associated HLA alleles or genes and those of the three other previously published HLA PheWAS studies (Figure 1a). Further, to evaluate the consistency of associations between the discovery and replication cohorts, we correlated the logistic regression log-odds ratios (betas) for the three types of analysis implemented here: HLA allele, diplotype and haplotype. Expectedly, we observed a strong correlation between the two independent cohorts (Pearson’s correlation coefficient about 0.9; Figure 1b).

**Figure 1.**
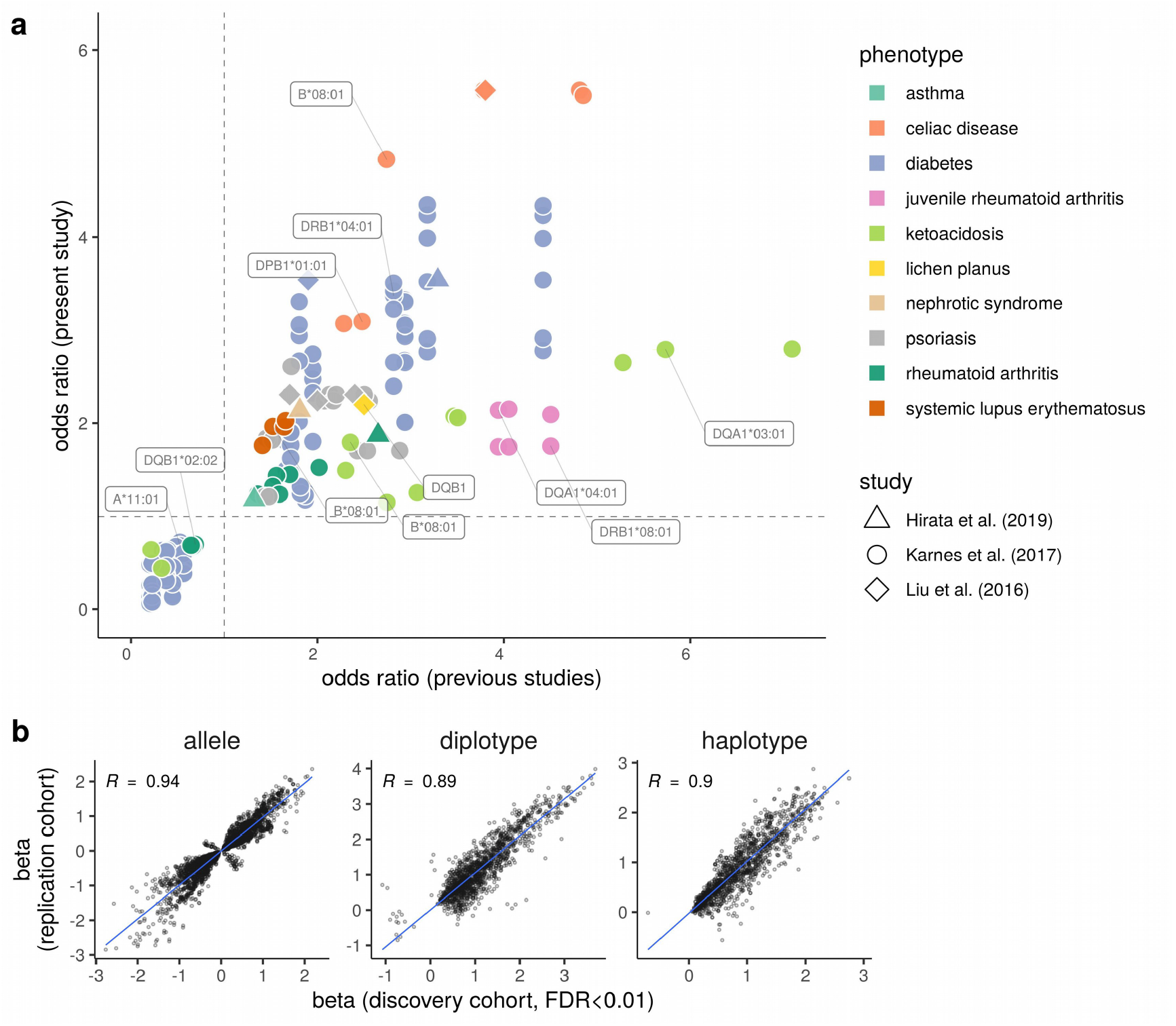
Comparison of HLA association effects. **a)** Odds ratios of previously reported HLA PheWAS associations (x-axis) vs. the discovery cohort of the present study (y-axis). Depending on the study, associations are shown either at the level of four-digit alleles (Karnes *et al*.) or at gene-level tagged by the highest ranking variant (Liu *et al*. & Hirata *et al*.). **b)** Correlation of HLA association FDR < 0.01 log-odds ratios (betas) between the discovery cohort (x-axis) and the replication cohort (y-axis) of the present study. Panels from left to right show the data for HLA allele, genotype, and two-locus haplotype association analyses.

We discovered statistically significant (discovery FDR < 0.01, replication p < 0.01) HLA allele associations in seven phenotypes for which we found scarce prior evidence of HLA association in the literature (Table 1). For example, we observed an association for *DQA1*\**01:03* and *DQB1*\**06:03* in mental and behavioural disorders due to cannabinoids (p-value = 10^−5^; beta = 0.6). Moreover, drug-induced hypoglycaemia without coma, vitreous haemorrhage, otitis externa, acute sinusitis, and trigger finger were all associated with *DQB1*\**03:02* and scleritis and episcleritis was associated with *B*\**27:05*.

**Table 1.** HLA allele associations and modifying effects in previously poorly studied phenotypes. Diplotype and haplotype analyses show effects of combinations of two alleles. Here, a strong protective effect on the risk allele can result in non-significant association.

| Type of analysis | Phenotype | Primary HLA | Modifying HLA | Discovery |  |  | Replication |  |
| --- | --- | --- | --- | --- | --- | --- | --- | --- |
|  |  |  |  | p-value | Beta | SE | Beta | SE |
| allele | Drug-induced hypoglycaemia without coma | DQB1*03:02 |  | 4.14E-05 | 0.667 | 0.155 | 0.917 | 0.173 |
|  |  | DQA1*03:01 |  | 4.83E-05 | 0.663 | 0.155 | 0.926 | 0.173 |
|  | Mental and behavioural disorders due to cannabinoids | DQA1*01:03 |  | 7.26E-06 | 0.603 | 0.128 | 0.439 | 0.127 |
|  |  | DQB1*06:03 |  | 1.22E-05 | 0.59 | 0.128 | 0.413 | 0.129 |
|  |  | DRB1*13:01 |  | 3.23E-05 | 0.54 | 0.125 | 0.378 | 0.125 |
|  | Vitreous haemorrhage | DQB1*03:02 |  | 1.36E-24 | 0.701 | 0.064 | 0.726 | 0.079 |
|  |  | DQA1*03:01 |  | 1.33E-23 | 0.688 | 0.065 | 0.707 | 0.079 |
|  |  | DRB1*04:01 |  | 4.61E-13 | 0.552 | 0.073 | 0.765 | 0.085 |
|  | Otitis externa | DQB1*03:02 |  | 1.39E-05 | 0.221 | 0.051 | 0.212 | 0.058 |
|  |  | B*18:01 |  | 2.46E-05 | 0.291 | 0.068 | 0.341 | 0.08 |
|  |  | DQA1*03:01 |  | 4.32E-05 | 0.209 | 0.051 | 0.208 | 0.059 |
|  | Acute sinusitis | DQA1*03:01 |  | 1.97E-07 | 0.147 | 0.028 | 0.145 | 0.033 |
|  |  | DQB1*03:02 |  | 2.66E-07 | 0.145 | 0.028 | 0.142 | 0.032 |
|  |  | DRB1*04:01 |  | 7.41E-06 | 0.142 | 0.032 | 0.134 | 0.036 |
|  | Trigger finger | DQB1*03:02 |  | 7.39E-08 | 0.333 | 0.061 | 0.261 | 0.074 |
|  |  | DQA1*03:01 |  | 7.67E-08 | 0.333 | 0.061 | 0.257 | 0.074 |
|  |  | DRB1*04:01 |  | 1.88E-06 | 0.328 | 0.068 | 0.357 | 0.079 |
|  | Scleritis and episcleritis | B*27:05 |  | 8.48E-08 | 0.579 | 0.102 | 0.574 | 0.122 |
| diplotype | Lichen planus | DQB1*05:01 | DQB1*05:01<sup>a</sup> | 2.46E-14 | 1.323 | 0.166 | 1.400 | 0.216 |
|  |  |  | DQB1*03:01 | 2.26E-02 | 0.385 | 0.165 | 0.621 | 0.201 |
|  | Seropositive rheumatoid arthritis | DRB1*04:08 | DRB1*04:01<sup>a</sup> | 5.84E-35 | 1.739 | 0.146 | 2.490 | 0.204 |
|  |  |  | DRB1*08:01 | 2.50E-01 | 0.205 | 0.223 | 0.226 | 0.384 |
|  | Co-morbidities, CVD and metabolic diseases | DRB1*04:01 | DRB1*03:01<sup>a</sup> | 1.27E-76 | 0.953 | 0.052 | 0.877 | 0.068 |
|  |  |  | DRB1*15:01 | 6.61E-01 | 0.020 | 0.045 | -0.067 | 0.061 |
|  |  |  | DRB1*11:01 | 9.20E-01 | 0.009 | 0.089 | 0.204 | 0.124 |
|  |  |  | DRB1*14:54 | 3.38E-01 | -0.167 | 0.177 | -0.026 | 0.24 |
|  | Thyroiditis, ILD-related definition | DQB1*02:01 | DQB1*03:02<sup>a</sup> | 4.71E-15 | 1.203 | 0.134 | 0.989 | 0.172 |
|  |  |  | DQB1*05:01 | 8.74E-03 | 0.387 | 0.144 | -0.112 | 0.207 |
| haplotype | Type1 diabetes, definitions combined | DQB1*03:02 | B*44:27<sup>a</sup> | 7.24E-14 | 2.223 | 0.255 | 2.190 | 0.309 |
|  |  |  | B*40:01 | 6.04E-08 | 0.885 | 0.15 | 1.129 | 0.165 |
|  |  |  | B*27:05 | 3.50E-06 | 0.823 | 0.164 | 0.713 | 0.214 |
|  | Diabetes, kidney failure | DQB1*03:02 | B*56:01<sup>a</sup> | 8.34E-19 | 1.333 | 0.129 | 1.240 | 0.177 |
|  |  |  | B*27:05 | 3.86E-03 | 0.518 | 0.173 | -0.355 | 0.366 |
|  |  |  | B*18:01 | 1.58E-02 | 0.371 | 0.149 | 0.486 | 0.19 |
|  | Diabetic maculopathy | DQB1*03:02 | B*56:01<sup>a</sup> | 1.34E-17 | 1.353 | 0.136 | 1.502 | 0.176 |
|  |  |  | B*18:01 | 4.83E-04 | 0.548 | 0.15 | 0.656 | 0.191 |
|  |  |  | B*27:05 | 3.70E-01 | 0.217 | 0.215 | -0.171 | 0.366 |
|  | ILD Co-morbidities, CVD and metabolic diseases | DRB1*04:01 | B*44:27<sup>a</sup> | 1.51E-06 | 0.695 | 0.147 | 0.586 | 0.18 |
|  |  |  | B*44:02 | 8.63E-03 | 0.100 | 0.038 | 0.042 | 0.052 |
|  | Other (seronegative) rheumatoid arthritis, wide | B*27:05 | DRB1*04:08<sup>a</sup> | 2.04E-19 | 1.046 | 0.102 | 0.978 | 0.132 |
|  |  |  | DRB1*04:04 | 8.75E-01 | 0.047 | 0.191 | 0.090 | 0.249 |

<sup>a</sup> Protective effect was determined relative to this allele combination in diplotype and haplotype analyses.

### Cross-phenotype HLA allele associations

To evaluate possible independence of an HLA association between two phenotypes, we conducted analyses by including a phenotype as an additional covariate in the regression models. We observed that altogether 68 HLA alleles showed evidence of independent association with two or more phenotype categories (Supplementary Table 2). To study shared HLA associations in autoimmune and infectious diseases, we narrowed down the results for these phenotypes to include only alleles that in conditional analyses showed evidence of associating with infectious diseases independently of at least one autoimmune disease. The results are summarized by Figure 2, showing the alleles, p-values, phenotypes and effect sizes of the associations. We found 12 alleles in five infectious and five autoimmune diseases that fulfilled the above criteria of association. Nine HLA alleles, eight of which appeared to be parts of *C*\**07:01 – B*\**08:01 – DRB1*\**03:01 – DQA1*\**05:01 – DQB1*\**02:01* and *DRB1*\**04:01 – DQA1*\**03:01 – DQB1*\**03:02* haplotypes, as well as *B*\**13:02*, predisposed to both autoimmune diseases and infections. Three alleles, all part of the *DRB1*\**13:01 – DQA1*\**01:03 – DQB1*\**06:03* haplotype, showed a lower frequency in cases. Altogether ten alleles associated with two or more infectious-autoimmune disease pairs.

**Figure 2.**
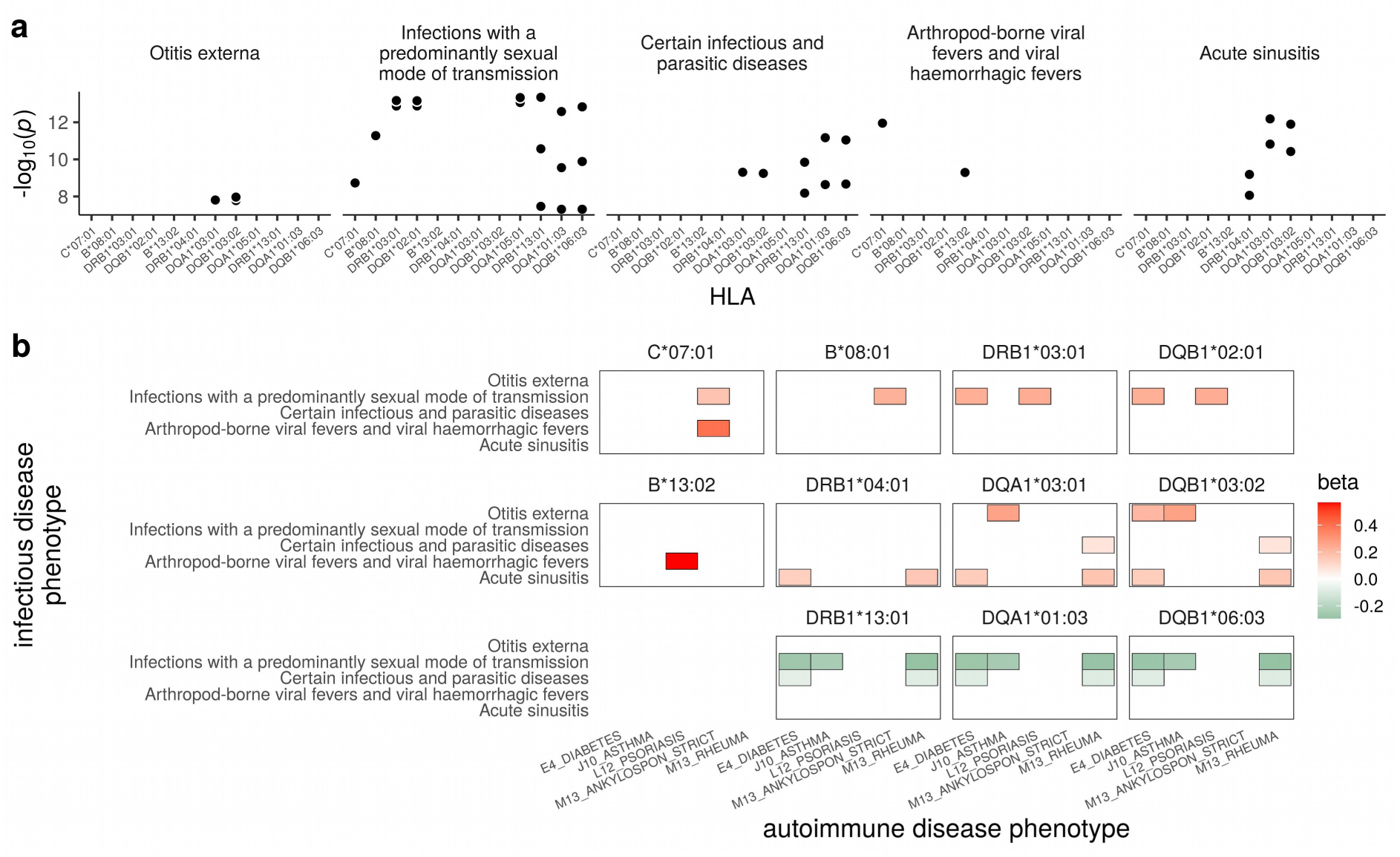
HLA alleles associated with an infectious disease independently of autoimmune disease. **a)** P-values of significant (< 5 × 10^−8^) alleles. Each panel show an infectious disease phenotype. Within panels, one allele can associate with more than one autoimmune disease. **b)** HLA alleles associated with infectious diseases (y-axis) independently from autoimmune diseases (x-axis). The color-filled squares indicate the effect size and direction. The results are grouped by known haplotypes in each row. DQA1^*^05:01 is omitted from the first row as its profile is indentical to the other two shown class II alleles. The data are based on conditional regression analyses with p < 5 × 10^−8^ threshold in the full dataset (discovery+replication), where selected phenotypes were analysed by adding a different phenotype as an additional covariate in the model one at a time.

### HLA diplotype associations

To analyze the effect of HLA risk allele diplotypes on the level of disease susceptibility, we conducted conditional regression analyses with diploid allele combinations. We found 225 statistically significant (discovery FDR < 0.01, replication p < 0.01) phenotypes representing 21 different phenotype categories associated with at least one risk allele diplotype (Supplementary Table 3). In 91 phenotypes representing 13 different phenotype categories the other HLA allele in the same locus exerted a statistically significant (discovery FDR < 0.01, replication p < 0.01) modifying effect on the risk allele (Supplementary Table 4). Figure 3 shows significant modifying allelic effects in phenotypes that deviated the most from expectation (i.e. the sum of individual allele effects, Figure 3a). For example, in type 1 diabetes, the results replicated the well-estalished protective allele *DQB1*\**06:02* and showed that *DQB1*\**04:02* increased the *DQB1*\**03:02* mediated risk for insulin medication despite having negative effect direction (−0.16) in the allele-level association analysis (Figure 3b). In coeliac disease, alleles such as *DQB1*\**06:03* or *DQB1*\**04:02*, that showed negative beta values in the single allele association test, contributed towards increasing the *DQB1*\**02:01* mediated risk (Figure 3b). Potentially novel heterozygotic effects on the risk allele are listed by Table 1.

**Figure 3.**
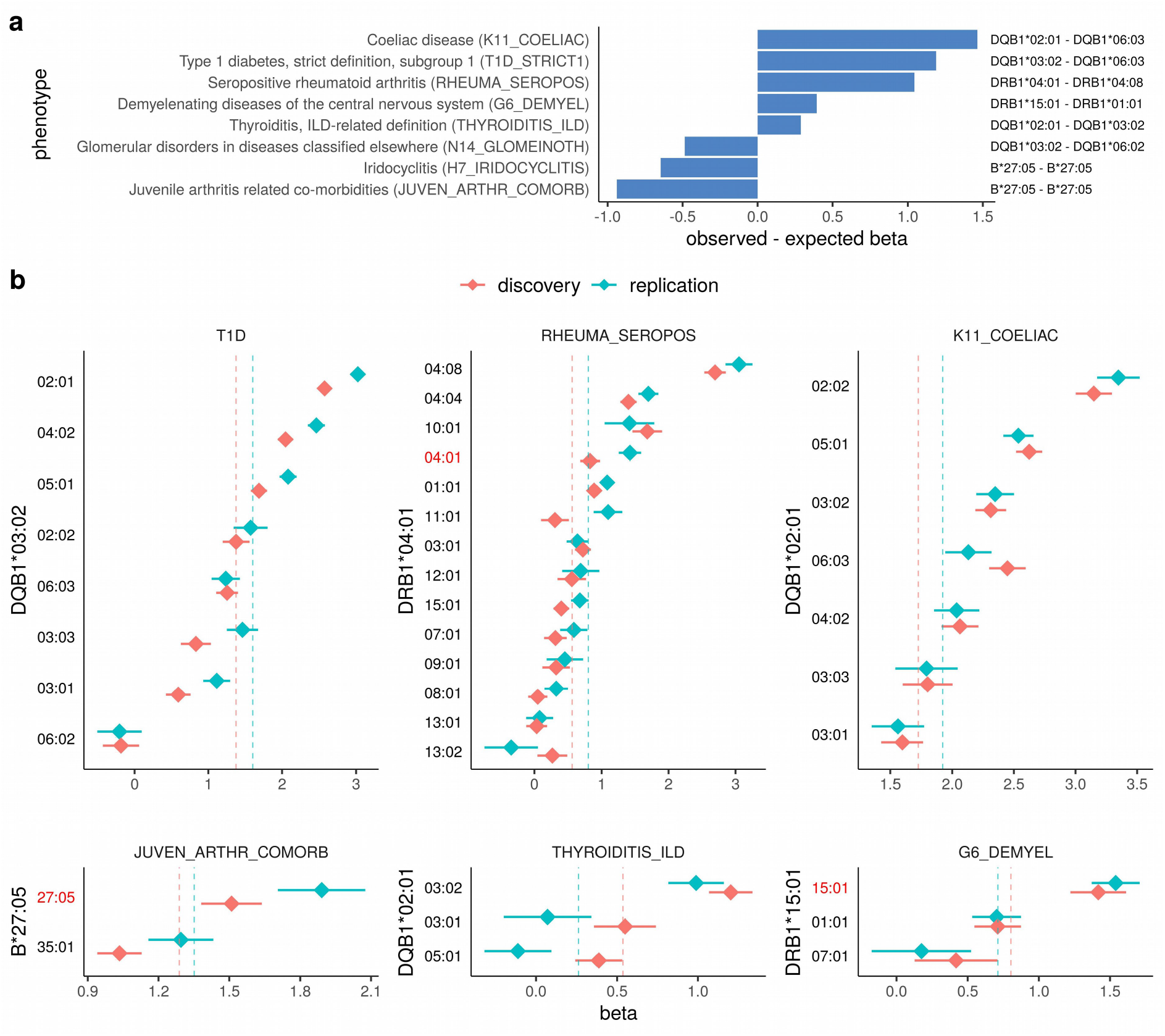
HLA diplotype effects. Risk allele in combination with the second allele of the same HLA gene. **a)** Phenotypes in which the risk allele diplotype association effects deviate from expected (i.e. sum of individual allelic effects). **b)** The x-axis shows log-odds ratios (betas) for different diplotypes depicted on the y-axis. The y-axis label indicates the primary risk allele, and the tick mark labels indicate the other alleles in the same locus. The vertical dashed lines indicate the risk allele’s effect estimates based on allele-level analysis. Only significant (discovery FDR < 0.01, replication p < 0.01) effects on the risk allele are shown. The error bars indicate standard errors for the beta values.

### HLA haplotype associations

To test whether the effect of a primary risk allele was affected by alleles of a HLA gene of a different class, we conducted conditional regression analyses with allele combinations from two HLA genes (termed here as haplotype associations). The analysis was performed using phased data but we cannot prove that they genuinely formed haplotypes. We found a total of 16 statistically significant haplotype associations with 224 phenotypes representing 23 different phenotype categories (Supplementary Table 5). There was a statistically significant (discovery FDR < 0.01, replication p < 0.01) modifying effect on the risk allele in 56 phenotypes representing 10 phenotype categories (Supplementary Table 6). Figure 4a shows significant modifying allelic effects in phenotypes that deviated the most from expectation (i.e. the sum of individual allele effects). For example, in T1D, even though *B*\**44:27* by itself was not associated, together with *DQB1*\**03:02* the risk is increased (Figure 4b). Potentially novel haplotype modifier effects on the risk allele are listed by Table 1.

**Figure 4.**
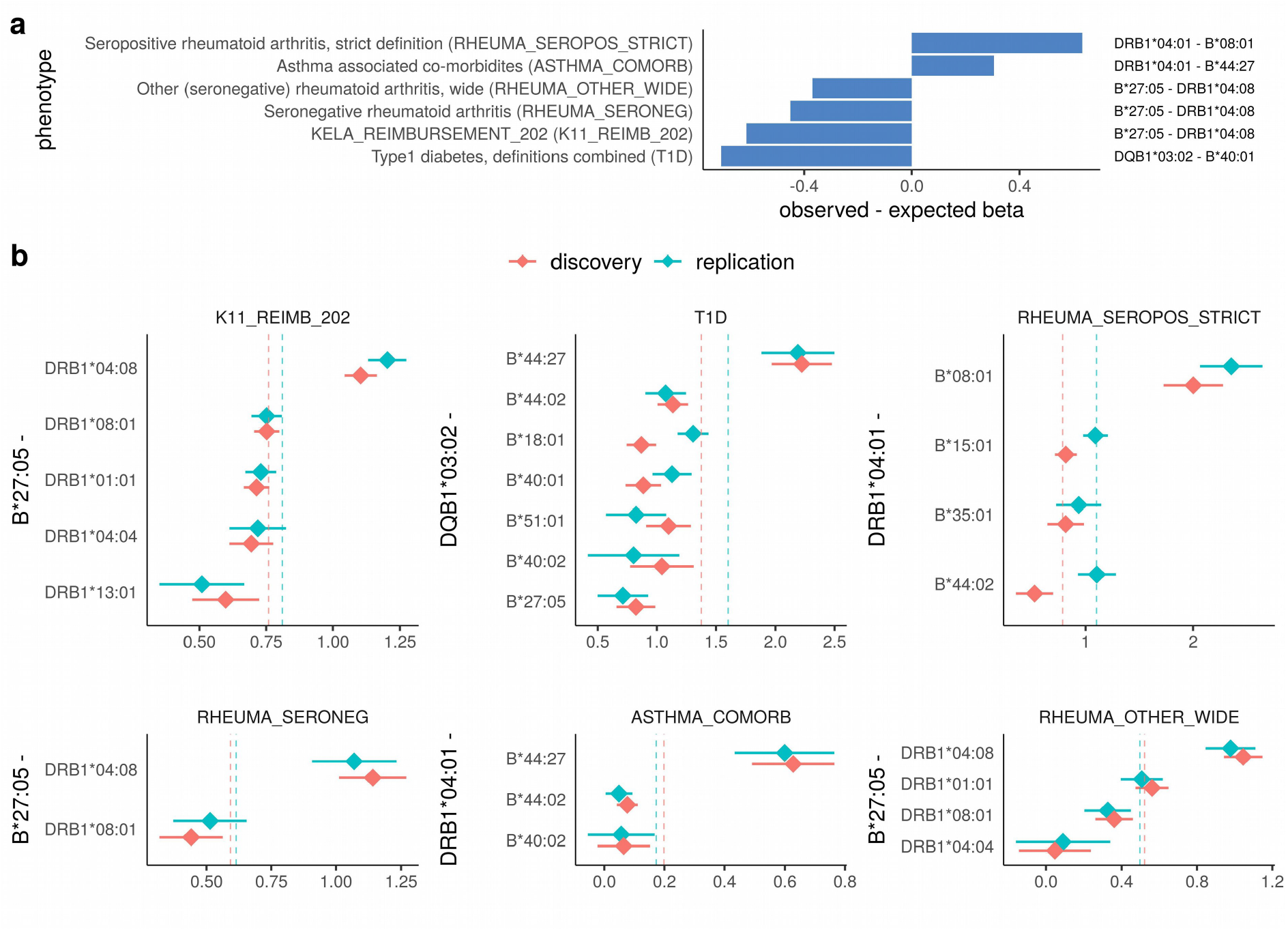
HLA haplotype effects. Risk allele in combination with another allele of a different HLA gene. **a)** Phenotypes in which the risk allele haplotype association effects deviate from expected (i.e. sum of individual allelic effects). **b)** The x-axis shows log-odds ratios (betas) for different two-locus allele combinations depicted on the y-axis. The y-axis label indicates the primary risk allele and the tick marks indicate alleles of a different HLA gene. The vertical dashed lines indicate the risk allele’s effect estimates based on allele-level analysis. Only significant (discovery FDR < 0.01, replication p < 0.01) effects on the risk allele are shown. The error bars indicate standard errors for the beta values.

## Discussion

The current study presents results of a systematic association analysis of imputed HLA alleles with over 3,000 clinical phenotypes in more than 235,000 individuals. In total, we report 3,649 statistically significant and successfully replicated allele-phenotype associations in 368 phenotypes distributed over 35 disease categories. Consistently with previous HLA PheWAS and other reports (Dendrou et al., 2018), our study uncovered well-established associations with major autoimmune disorders, and also found evidence of HLA pleiotropy (Karnes et al., 2017; Liu et al., 2016) in particular between infectious and autoimmune diseases. Expectedly, the effect size estimates between the previous studies and our discovery and replication data sets showed overall high concordance, validating the accuracy of HLA imputation, phenotype data and association analyses based on these. The results from conditional analyses focusing on selected combinations of HLA alleles and cross-phenotype associations further add to the existing knowledge by including risk-modifying effects not studied before in a phenome-wide context.

In a recent well-powered association study, MHC region was linked with multiple common infectious diseases, and fine-mapping revealed several independent signals among HLA-gene variants and alleles (Tian et al., 2017). Moreover, in another study on MHC expression quantitative trait loci, protection from bacterial infections in cystic fibrosis by the common autoimmune risk haplotype AH 8.1 was found to be mediated by a non-HLA gene carried in the same haplotype (D’Antonio et al., 2019). Our finding that certain HLA alleles in common haplotypes were shared by infectious and autoimmune diseases is intriguing in regard to the proposed triggering role of infections in autoimmunity (Ercolini and Miller, 2009). The result on the *B*\**13:02 – DQB1*\**03:02* and C\**07:01 – DQB1*\**02:01* haplotypes showed that class I and II alleles exhibited different associated phenotypes, suggesting that these alleles may have effects that are not explained by linkage disequilibrium alone. As evidence of HLA pleiotropy was also reported by two previous MHC PheWASs (Karnes et al., 2017; Liu et al., 2016), it will be of great interest to try to reveal the mechanistic background for these shared associations, especially between infections and autoimmunity (D’Antonio et al., 2019; Matzaraki et al., 2017).

The strong enrichment of HLA risk alleles in autoimmune diseases, e.g. DQ8 in T1D, DQ2 in coeliac disease, or B27 in arthropathies, automatically leads to lower frequencies of other alleles in the risk locus and consequently to risk-reducing effect estimates irrespective of actual association. Conditional analyses adjusted for allelic variation can reveal genuine effects of the risk-gene HLA genotypes. In line with previous analyses, our HLA diplotype PheWAS replicated known protective allelic effects in e.g. in demyelinating diseases (*DRB1*\**07:01* and *01:01*) (Wu et al., 2010), arthropathic psoriasis (*C*\**07:01*) (Queiro et al., 2006), diabetes (*DQB1*\**06:02*) (Pugliese et al., 1995), and seropositive RA (*DRB1:13:0*1 and *08:01*) (van der Helm-van Mil et al., 2005) and provided estimates for risk-modifying effects of a range of alleles occurring together with the top risk allele in autoimmune disorders. Our results showed a risk-modifying effect of *DQB1*\**03:01* for *DQB1*\**05:01* in lichen planus (LP), helping resolve the somewhat contradictry results obtained by previous serotyping studies on frequencies of DQ1 and DQ3 in LP patients (Nasa et al., 1995; Porter et al., 1993).

Population founder effect can lead to reduced genetic diversity and altered frequencies of genetic variants (Chheda et al., 2017), including HLA alleles and haplotypes (Creary et al., 2019; Hurley et al., 2020). The current study was based on genetically defined cohort of Finns that constitute a Northern European genetic isolate. A characteristic genetic architecture is visible in the repertoire of HLA haplotypes where a number of Finnish enriched rare (FER) haplotypes are substantially more common than elsewhere in Europe (Linjama et al., 2018). Our HLA class I – class II analysis demonstrates how haplotype effects can be estimated in a genetically characteristic population. We found that *B*\**27:05* occurring together with *DRB1*\**04:08* carried the highest risk for seronegative rheumatic diseases, confirming an association that has been previously described in the Finnish population (Tuokko et al., 1997). This allele combination occurs in *C*\**01:02 - HLA-B*\**27:05 – DRB1*\**04:08 – DQB1*\**03:01* haplotype that belongs to the FER group and is 3300 times more frequent in the Finnish population than in other European populations. Our study further demonstrated that the predisposing effect of *B*\**27:05* was effectively removed by *DRB1*\**04:04*. This allele pair is known to occur in the *C*\**01:02/02:02 – B*\**27:05 – DRB1*\**04:04 – DQB1*\**03:02* FER haplotype.

While HLA class I and II have been reported to be independently associated with T1D (Eike et al., 2009; Mikk et al., 2017), the compund effect of allelic heterogeneity between HLA class I and II remains less comprehensively understood. We observed protective effects for HLA class I alleles that by themselves did not have association with T1D and its comorbidities in our analyses or elsewhere in the literature (Noble and Valdes, 2011). For example, *B*\**27:05* and B^*^*40:01* occurring together with *DQB1*\**03:02* reduced the risk conferred by *DQB1*\**03:02* while *B*\**44:27* substantially increased it. The predisposing effect of the uncommon *B*\**44:27* allele in diabetes-related conditions can go unnoticed in mixed populations due its infrequency or appearance in different class II haplotypes. Allele *B*\**44:27* is relatively rare also in Finland and occurs mostly with *C*\**07:04 – B*\**44:27 – DRB1*\**16:01 – DQB1*\**05:02, DRB1*\**08:01 – DQB1*\**04:02* and *DRB1*\**01:01 – DQB1*\**05:01*. As these haplotypes lack known risk alleles, the causative variant remains unknown but suggests a potential role for *B*\**44:27*. Obviously, rare alleles such as *B*\**44:27* and haplotypes carrying it are not widely studied, and also the risk factor associated with *B*\**44:27* may not be the same as in *DQB1*\**03:02* haplotypes.

Our study is also limited in some respects. First, analysis of HLA alleles alone cannot definitively attribute the observed associations directly to HLA owing to strong linkage disequilibrium within the MHC (Trowsdale and Knight, 2013). For example, the known associations between disorders of iron metabolism and *A*\**03:01*, and that between disorders of adrenal gland and *DRB1*\**04:04*, at least partially are a result from linkage disequilibrium with *HFE* gene and *CYP21* gene, respectively. Also, most of the rare HLA alleles were not covered by the used imputation panel and consequently the analysis did not cover their possible associations. Second, our study is restricted by statistical power particularly in conditional analyses with many covariates and in endpoints having a low number of cases. While the independent replication design of the study helps eliminate non-systematical false positives arising from e.g. relatedness, batch and other chance factors, it cannot categorically rule them out or remove sampling uncertainty in low-powered endpoints. Third, the FinnGen phenotypes, albeit carefully curated, were derived from health register which cannot be assumed to be totally accurate. Finally, haplotype analysis cannot prove that the alleles are encoded in *cis*, but the effects between two HLA genes, or chromosomal regions between them, can also take place in *trans*.

In conclusion, the results of the present study illustrate the role of HLA alleles both separately and in combination in immune-mediated diseases, revealing potentially new HLA-linked disease phenotypes and providing a data resource for future HLA analyses in independent populations. The results expand the view of the complex genetic structure of HLA, motivating the consideration of allele and gene interactions in risk calculations. These results can serve as starting points for functional studies focusing on mechanistic molecular underpinnings of the discovered associations.

## Materials and Methods

### Subjects and clinical endpoints

The discovery cohort of the study included all biobank participants in the FinnGen (www.finngen.fi) data release R3 (n_total_ = 146,630) while the independent replication cohort comprised the data release R5 (without R3; n_total_ = 89,340). Numbers of cases and controls for each phenotype are given in the Supplementary Table 1. Endpoints with at least 5 cases carrying a given allele in both discovery and replication cohorts were included in the analysis. The clinical disease endpoint definitions were curated from ICD 9-10, ICD-O-3, the Social Insurance Institute (KELA) drug reimbursement codes and ATC-codes as a part of the FinnGen project (finngen.gitbook.io/documentation/methods/endpoints). For clarity, the FinnGen phenotypes include many partially overlapping diseases or traits, particularly in diabetes and its comorbidities. Thus, the included phenotypes are not necessarily independent. All patients and control subjects provided an informed consent for biobank research in accordance with the Finnish Biobank Act, with the exception of FinnGen legacy samples which were approved by the National Supervisory Authority for Welfare and Health (Valvira). The FinnGen study protocol was approved by the Ethical Review Board of the Hospital District of Helsinki and Uusimaa (Nr HUS/990/2017). All samples and individual-level data were pseudonymized and processed in accordance with the EU GDPR law.

### Genotyping

Genotyping of FinnGen samples was performed on a customized ThermoFisher Axiom array at the Thermo Fisher genotyping service facility (San Diego, USA). Genotype calling and quality control steps are described in finngen.gitbook.io/documentation/methods/genotype-imputation. The array markes files can be downloaded from www.finngen.fi/en/researchers/genotyping. The protocol for genotype liftover to hg38/GRCh38 is described in detail in www.protocols.io/view/genotyping-chip-data-lift-over-to-reference-genome-xbhfij6?version_warning=no, and genotype imputation protocol is described in www.protocols.io/view/genotype-imputation-workflow-v3-0-xbgfijw.

### HLA allele analysis

We implemented the PheWAS approach (Denny et al., 2013) for imputed alleles of *HLA-A*, -*B*, -*C*, -*DRB1*, -*DQA1*, -*DQB1* and -*DPB1* genes to analyze their correlation with 3,355 clinical case-control endpoints in 37 broad disease categories. Each analysed phenotype included at least five cases in both discovery and replication sets. HLA imputation at four-digit resolution (i.e. protein-level) was conducted as described previously (Ritari et al., 2020). Briefly, we used HIBAG v1.18.1 (Zheng et al., 2014) R library with a Finnish population-specific HLA reference panel (n = 1,150) based on ∼4,500 SNPs within the MHC region (chr6:28.51-33.48 Mb; hg38/GRCh38), and considered imputation posterior probabilities > 0.5 as acceptable. For association analyses, we defined the imputed HLA alleles as bi-allelic SNPs and assumed additive effects of allele dosages on the binary phenotype. Logistic regression models were run using SPAtest v3.0.2 (Dey et al., 2017) in R v3.6.3 (R Core Team, 2020) with top 10 genetic principal components (PCs), age and sex as covariates. To correct for multiple testing under dependency and to identify associations for validation in the replication cohort, we applied adaptive Benjamini-Hochberg (Benjamini and Hochberg, 2000; Kim and van de Wiel, 2008) procedure to the discovery cohort SPAtest saddlepoint approximated p-values using the R library mutoss v0.1-12 (MuToss Coding Team et al., 2017) at FDR < 0.01 threshold. We considered an association valid if the replication p-value was < 0.01 and the effect direction was consistent with the discovery cohort.

To evaluate independent contributions of HLA alleles significantly associated with multiple disease categories, we performed conditional analyses that systematically included a phenotype from a different disease category as an additional covariate. In this analysis we used the whole dataset (data release R5) and genome-wide p-value threshold of 5×10^−8^. To exclude phenotypes in strong correlation with each other from the analysis, we first computed an all-vs-all Pearson’s correlation matrix between the phenotypes and removed those having a correlation >0.8 with another phenotype. Association for each HLA allele with a given phenotype was performed by including a different, non-correlating phenotype as a covariate along with age, sex and 10 genetic PCs using SPAtest as described above.

### HLA diplotype analysis

To systematically study how the association effect of the primary risk allele was impacted by other alleles of the same HLA gene, we performed association analyses for HLA allele combinations (termed here as diplotypes). The top risk alleles were identified based on the lowest significant single-allele p-value for each phenotype in the discovery cohort. We performed conditional regression analyses by including all the diplotypes in the same model for a given phenotype. With this approach our aim was to quantify actual allelic effects as distinguished from nonpredisposition to the risk allele. As described above, the top 10 genetic PCs, age and sex were included as other covariates. To identify significant effects relative to the top risk genotype for a given phenotype, we performed a two-tailed Z-test on the obtained conditional logistic regression coefficients (betas) and their standard errors.

### HLA haplotype analysis

The haplotype analysis was based on the observation that in some phenotypes a significant association was found both in HLA class I and class II genes. To evaluate whether alleles in a class I gene affected the risk of an allele in a class II gene, or vice versa, we considered combinations of alleles from both class I and II. The top risk allele for each phenotype was first identified based on the lowest significant single-allele p-value in the discovery cohort, and then combined with alleles of a HLA gene of a different class (termed here as haplotypes). Thus, the primary risk allele was studied in all available allele combinations of the secondary gene. HLAs were imputed on phased genotype data obtained from genotype imputation, and the combined loci under analysis were selected from the same phase. All haplotypes were included in the same regression model for a given phenotype. Two-tailed Z-test was used to evaluate the significance of the haplotoypic effects.

### Data availability

The FinnGen summary statistics data can be accessed through the Finnish Biobanks’ FinnBB portal and the FinnGen website (https://www.finngen.fi/en/access_results).

### Code availability

The analysis code is available at https://github.com/FRCBS/HLA_PheWAS. The FinnGen genotyping and imputation protocol is described at https://doi-org.libproxy.helsinki.fi/10.17504/protocols.io.nmndc5e.

## Data Availability

The FinnGen summary statistics data can be accessed through the Finnish
Biobanks' FinnBB portal (www.finbb.fi).

## Acknowledgements

The study was supported by the Academy of Finland, the Finnish Cancer Association, VTR funding from the Finnish Government, and Business Finland. FinnGen funding statement is available as supplementary information. We are grateful to all FinnGen participants for their generous contribution to the project. The funders and biobanks had no role in study design, data collection and analysis, decision to publish, or preparation of the manuscript.

## References

A. Barr T, Gray M, Gray D. 2012. B Cells: Programmers of CD4 T Cell Responses. IDDT 12:222–231. doi:10.2174/187152612800564446

Bach J-F. 2018. The hygiene hypothesis in autoimmunity: the role of pathogens and commensals. Nat Rev Immunol 18:105–120. doi:10.1038/nri.2017.111

Benjamini Y, Hochberg Y. 2000. On the Adaptive Control of the False Discovery Rate in Multiple Testing With Independent Statistics. Journal of Educational and Behavioral Statistics 25:60–83. doi:10.3102/10769986025001060

Bettencourt A, Carvalho C, Leal B, Brás S, Lopes D, Martins da Silva A, Santos E, Torres T, Almeida I, Farinha F, Barbosa P, Marinho A, Selores M, Correia J, Vasconcelos C, Costa PP, da Silva BM. 2015. The Protective Role of HLA-DRB1 13 in Autoimmune Diseases. Journal of Immunology Research 2015:1–6. doi:10.1155/2015/948723

Chheda H, Palta P, Pirinen M, McCarthy S, Walter K, Koskinen S, Salomaa V, Daly M, Durbin R, Palotie A, Aittokallio T, Ripatti S. 2017. Whole-genome view of the consequences of a population bottleneck using 2926 genome sequences from Finland and United Kingdom. Eur J Hum Genet 25:477– 484. doi:10.1038/ejhg.2016.205

Creary LE, Gangavarapu S, Mallempati KC, Montero-Martín G, Caillier SJ, Santaniello A, Hollenbach JA, Oksenberg JR, Fernández-Viña MA. 2019. Next-generation sequencing reveals new information about HLA allele and haplotype diversity in a large European American population. Human Immunology 80:807–822. doi:10.1016/j.humimm.2019.07.275

D’Antonio M, Reyna J, Jakubosky D, Donovan MK, Bonder M-J, Matsui H, Stegle O, Nariai N, D’Antonio-Chronowska A, Frazer KA. 2019. Systematic genetic analysis of the MHC region reveals mechanistic underpinnings of HLA type associations with disease. eLife 8:e48476. doi:10.7554/eLife.48476

Dendrou CA, Petersen J, Rossjohn J, Fugger L. 2018. HLA variation and disease. Nat Rev Immunol 18:325–339. doi:10.1038/nri.2017.143

Denny JC, Bastarache L, Ritchie MD, Carroll RJ, Zink R, Mosley JD, Field JR, Pulley JM, Ramirez AH, Bowton E, Basford MA, Carrell DS, Peissig PL, Kho AN, Pacheco JA, Rasmussen LV, Crosslin DR, Crane PK, Pathak J, Bielinski SJ, Pendergrass SA, Xu H, Hindorff LA, Li R, Manolio TA, Chute CG, Chisholm RL, Larson EB, Jarvik GP, Brilliant MH, McCarty CA, Kullo IJ, Haines JL, Crawford DC, Masys DR, Roden DM. 2013. Systematic comparison of phenome-wide association study of electronic medical record data and genome-wide association study data. Nat Biotechnol 31:1102–1111. doi:10.1038/nbt.2749

Dey R, Schmidt EM, Abecasis GR, Lee S. 2017. A Fast and Accurate Algorithm to Test for Binary Phenotypes and Its Application to PheWAS. The American Journal of Human Genetics 101:37–49. doi:10.1016/j.ajhg.2017.05.014

Diogo D, Tian C, Franklin CS, Alanne-Kinnunen M, March M, Spencer CCA, Vangjeli C, Weale ME, Mattsson H, Kilpeläinen E, Sleiman PMA, Reilly DF, McElwee J, Maranville JC, Chatterjee AK, Bhandari A, Nguyen K-DH, Estrada K, Reeve M-P, Hutz J, Bing N, John S, MacArthur DG, Salomaa V, Ripatti S, Hakonarson H, Daly MJ, Palotie A, Hinds DA, Donnelly P, Fox CS, Day-Williams AG, Plenge RM, Runz H. 2018. Phenome-wide association studies across large population cohorts support drug target validation. Nat Commun 9:4285. doi:10.1038/s41467-018-06540-3

Eike MC, Becker T, Humphreys K, Olsson M, Lie BA. 2009. Conditional analyses on the T1DGC MHC dataset: novel associations with type 1 diabetes around HLA-G and confirmation of HLA-B. Genes Immun 10:56–67. doi:10.1038/gene.2008.74

Ercolini AM, Miller SD. 2009. The role of infections in autoimmune disease. Clinical & Experimental Immunology 155:1–15. doi:10.1111/j.1365-2249.2008.03834.x

Furukawa H, Oka S, Tsuchiya N, Shimada K, Hashimoto A, Tohma S, Kawasaki A. 2017. The role of common protective alleles HLA-DRB1*13 among systemic autoimmune diseases. Genes Immun 18:1–7. doi:10.1038/gene.2016.40

Gregersen PK, Silver J, Winchester RJ. 1987. The shared epitope hypothesis. an approach to understanding the molecular genetics of susceptibility to rheumatoid arthritis. Arthritis & Rheumatism 30:1205–1213. doi:10.1002/art.1780301102

Hirata J, Hosomichi K, Sakaue S, Kanai M, Nakaoka H, Ishigaki K, Suzuki K, Akiyama M, Kishikawa T, Ogawa K, Masuda T, Yamamoto K, Hirata M, Matsuda K, Momozawa Y, Inoue I, Kubo M, Kamatani Y, Okada Y. 2019. Genetic and phenotypic landscape of the major histocompatibilty complex region in the Japanese population. Nat Genet 51:470–480. doi:10.1038/s41588-018-0336-0

Hu X, Deutsch AJ, Lenz TL, Onengut-Gumuscu S, Han B, Chen W-M, Howson JMM, Todd JA, de Bakker PIW, Rich SS, Raychaudhuri S. 2015. Additive and interaction effects at three amino acid positions in HLA-DQ and HLA-DR molecules drive type 1 diabetes risk. Nat Genet 47:898–905. doi:10.1038/ng.3353

Hurley CK, Kempenich J, Wadsworth K, Sauter J, Hofmann JA, Schefzyk D, Schmidt AH, Galarza P, Cardozo MBR, Dudkiewicz M, Houdova L, Jindra P, Sorensen BS, Jagannathan L, Mathur A, Linjama T, Torosian T, Freudenberger R, Manolis A, Mavrommatis J, Cereb N, Manor S, Shriki N, Sacchi N, Ameen R, Fisher R, Dunckley H, Andersen I, Alaskar A, Alzahrani M, Hajeer A, Jawdat D, Nicoloso G, Kupatawintu P, Cho L, Kaur A, Bengtsson M, Dehn J. 2020. Common, intermediate and well-documented HLA alleles in world populations: CIWD version 3.0.0. HLA 95:516–531. doi:10.1111/tan.13811

Karnes JH, Bastarache L, Shaffer CM, Gaudieri S, Xu Y, Glazer AM, Mosley JD, Zhao S, Raychaudhuri S, Mallal S, Ye Z, Mayer JG, Brilliant MH, Hebbring SJ, Roden DM, Phillips EJ, Denny JC. 2017. Phenome-wide scanning identifies multiple diseases and disease severity phenotypes associated with HLA variants. Sci Transl Med 9:eaai8708. doi:10.1126/scitranslmed.aai8708

Kim KI, van de Wiel MA. 2008. Effects of dependence in high-dimensional multiple testing problems. BMC Bioinformatics 9:114. doi:10.1186/1471-2105-9-114

Lenz TL, Deutsch AJ, Han B, Hu X, Okada Y, Eyre S, Knapp M, Zhernakova A, Huizinga TWJ, Abecasis G, Becker J, Boeckxstaens GE, Chen W-M, Franke A, Gladman DD, Gockel I, Gutierrez-Achury J, Martin J, Nair RP, Nöthen MM, Onengut-Gumuscu S, Rahman P, Rantapää-Dahlqvist S, Stuart PE, Tsoi LC, van Heel DA, Worthington J, Wouters MM, Klareskog L, Elder JT, Gregersen PK, Schumacher J, Rich SS, Wijmenga C, Sunyaev SR, de Bakker PIW, Raychaudhuri S. 2015. Widespread non-additive and interaction effects within HLA loci modulate the risk of autoimmune diseases. Nat Genet 47:1085–1090. doi:10.1038/ng.3379

Linjama T, Eberhard H-P, Peräsaari J, Müller C, Korhonen M. 2018. A European HLA Isolate and Its Implications for Hematopoietic Stem Cell Transplant Donor Procurement. Biology of Blood and Marrow Transplantation 24:587– 593. doi:10.1016/j.bbmt.2017.10.010

Liu J, Ye Z, Mayer JG, Hoch BA, Green C, Rolak L, Cold C, Khor S-S, Zheng X, Miyagawa T, Tokunaga K, Brilliant MH, Hebbring SJ. 2016. Phenome-wide association study maps new diseases to the human major histocompatibility complex region. J Med Genet 53:681–689. doi:10.1136/jmedgenet-2016-103867

Matzaraki V, Kumar V, Wijmenga C, Zhernakova A. 2017. The MHC locus and genetic susceptibility to autoimmune and infectious diseases. Genome Biol 18:76. doi:10.1186/s13059-017-1207-1

Mikk M-L, Heikkinen T, El-Amir MI, Kiviniemi M, Laine A-P, Härkönen T, Veijola R, Toppari J, Knip M, Ilonen J, the Finnish Paediatric Diabetes Register. 2017. The association of the HLA-A*24:02, B*39:01 and B*39:06 alleles with type 1 diabetes is restricted to specific HLA-DR/DQ haplotypes in Finns. HLA 89:215–224. doi:10.1111/tan.12967

MuToss Coding Team, Blanchard G, Dickhaus T, Hack N, Konietschke F, Rohmeyer K, Rosenblatt J, Scheer M, Werft W. 2017. mutoss: Unified Multiple Testing Procedures.

Nasa GL, Cottoni F, Mulargia M, Carcassi C, Vacca A, Pizzati A, Ledda A, Montesu MA, Cerimele D, Contu L. 1995. HLA antigen distribution in different clinical subgroups demonstrates genetic heterogeneity in lichen planus. British Journal of Dermatology 132:897–900. doi:10.1111/j.1365-2133.1995.tb16945.x

Noble JA, Valdes AM. 2011. Genetics of the HLA Region in the Prediction of Type 1 Diabetes. Curr Diab Rep 11:533–542. doi:10.1007/s11892-011-0223-x

Okada Y, Suzuki A, Ikari K, Terao C, Kochi Y, Ohmura K, Higasa K, Akiyama M, Ashikawa K, Kanai M, Hirata J, Suita N, Teo Y-Y, Xu H, Bae S-C, Takahashi A, Momozawa Y, Matsuda K, Momohara S, Taniguchi A, Yamada R, Mimori T, Kubo M, Brown MA, Raychaudhuri S, Matsuda F, Yamanaka H, Kamatani Y, Yamamoto K. 2016. Contribution of a Non-classical HLA Gene, HLA-DOA, to the Risk of Rheumatoid Arthritis. The American Journal of Human Genetics 99:366–374. doi:10.1016/j.ajhg.2016.06.019

Oldstone MBA. 1998. Molecular mimicry and immune-mediated diseases. FASEB j 12:1255–1265. doi:10.1096/fasebj.12.13.1255

Porter K, Klouda P, Scully C, Bidwell J, Porter S. 1993. Class I and II HLA antigens in British patients with oral lichen planus. Oral Surgery, Oral Medicine, Oral Pathology 75:176–180. doi:10.1016/0030-4220(93)90090-Q

Pugliese A, Gianani R, Moromisato R, Awdeh ZL, Alper CA, Erlich HA, Jackson RA, Eisenbarth GS. 1995. HLA-DQB1*0602 Is Associated With Dominant Protection From Diabetes Even Among Islet Cell Antibody-Positive First-Degree Relatives of Patients with IDDM. Diabetes 44:608–613. doi:10.2337/diab.44.6.608

Queiro R, Gonzalez S, López-Larrea C, Alperi M, Sarasqueta C, Riestra J, Ballina J. 2006. HLA-C locus alleles may modulate the clinical expression of psoriatic arthritis. Arthritis Res Ther 8:R185. doi:10.1186/ar2097

R Core Team. 2020. R: A language and environment for statistical computing. Vienna, Austria: R Foundation for Statistical Computing.

Raychaudhuri S, Sandor C, Stahl EA, Freudenberg J, Lee H-S, Jia X, Alfredsson L, Padyukov L, Klareskog L, Worthington J, Siminovitch KA, Bae S-C, Pleng RM, Gregersen PK, de Bakker PIW. 2012. Five amino acids in three HLA proteins explain most of the association between MHC and seropositive rheumatoid arthritis. Nat Genet 44:291–296. doi:10.1038/ng.1076

Ritari J, Hyvärinen K, Clancy J FinnGen, Partanen J, Koskela S. 2020. Increasing accuracy of HLA imputation by a population-specific reference panel in a FinnGen biobank cohort. NAR Genomics and Bioinformatics 2:lqaa030. doi:10.1093/nargab/lqaa030

Sfriso P, Ghirardello A, Botsios C, Tonon M, Zen M, Bassi N, Bassetto F, Doria A. 2010. Infections and autoimmunity: the multifaceted relationship. Journal of Leukocyte Biology 87:385–395. doi:10.1189/jlb.0709517

The International HIV Controllers Study. 2010. The Major Genetic Determinants of HIV-1 Control Affect HLA Class I Peptide Presentation. Science 330:1551– 1557. doi:10.1126/science.1195271

The MHC sequencing consortium. 1999. Complete sequence and gene map of a human major histocompatibility complex. Nature 401:921–923. doi:10.1038/44853

Thorsby E. 2009. A short history of HLA. Tissue Antigens 74:101–116. doi:10.1111/j.1399-0039.2009.01291.x

Tian C, Hromatka BS, Kiefer AK, Eriksson N, Noble SM, Tung JY, Hinds DA. 2017. Genome-wide association and HLA region fine-mapping studies identify susceptibility loci for multiple common infections. Nat Commun 8:599. doi:10.1038/s41467-017-00257-5

Trowsdale J, Knight JC. 2013. Major Histocompatibility Complex Genomics and Human Disease. Annu Rev Genom Hum Genet 14:301–323. doi:10.1146/annurev-genom-091212-153455

Tsai S, Santamaria P. 2013. MHC Class II Polymorphisms, Autoreactive T-Cells, and Autoimmunity. Front Immunol 4. doi:10.3389/fimmu.2013.00321

Tuokko J, Reijonen H, Ilonen J, Anttila K, Nikkari S, Mottonen T, Yli-Kerttula U, Toivanen A. 1997. Increase of HLA-DRB1*0408 and -DQB1*0301 in HLA-B27 positive reactive arthritis. Annals of the Rheumatic Diseases 56:37– 40. doi:10.1136/ard.56.1.37

van der Helm-van Mil AHM, Huizinga TWJ, Schreuder GMTh, Breedveld FC, de Vries RRP, Toes REM. 2005. An independent role of protective HLA class II alleles in rheumatoid arthritis severity and susceptibility. Arthritis Rheum 52:2637–2644. doi:10.1002/art.21272

van Lummel M, Buis DTP, Ringeling C, de Ru AH, Pool J, Papadopoulos GK, van Veelen PA, Reijonen H, Drijfhout JW, Roep BO. 2019. Epitope Stealing as a Mechanism of Dominant Protection by HLA-DQ6 in Type 1 Diabetes. Diabetes 68:787–795. doi:10.2337/db18-0501

Verma A, Lucas A, Verma SS, Zhang Y, Josyula N, Khan A, Hartzel DN, Lavage DR, Leader J, Ritchie MD, Pendergrass SA. 2018. PheWAS and Beyond: The Landscape of Associations with Medical Diagnoses and Clinical Measures across 38,662 Individuals from Geisinger. The American Journal of Human Genetics 102:592–608. doi:10.1016/j.ajhg.2018.02.017

Wu J-S, James I, Wei Qiu, Castley A, Christiansen FT, Carroll WM, Mastaglia FL, Kermode AG. 2010. Influence of HLA-DRB1 allele heterogeneity on disease risk and clinical course in a West Australian MS cohort: a high-resolution genotyping study. Mult Scler 16:526–532. doi:10.1177/1352458510362997

Wucherpfennig KW, Strominger JL. 1995. Molecular mimicry in T cell-mediated autoimmunity: Viral peptides activate human T cell clones specific for myelin basic protein. Cell 80:695–705. doi:10.1016/0092-8674(95)90348-8

Zheng X, Shen J, Cox C, Wakefield JC, Ehm MG, Nelson MR, Weir BS. 2014. HIBAG —HLA genotype imputation with attribute bagging. Pharmacogenomics J 14:192–200. doi:10.1038/tpj.2013.18

