## Supplementary figures and text. for "Phenome-wide HLA association landscape of 235,000 Finnish biobank participants"

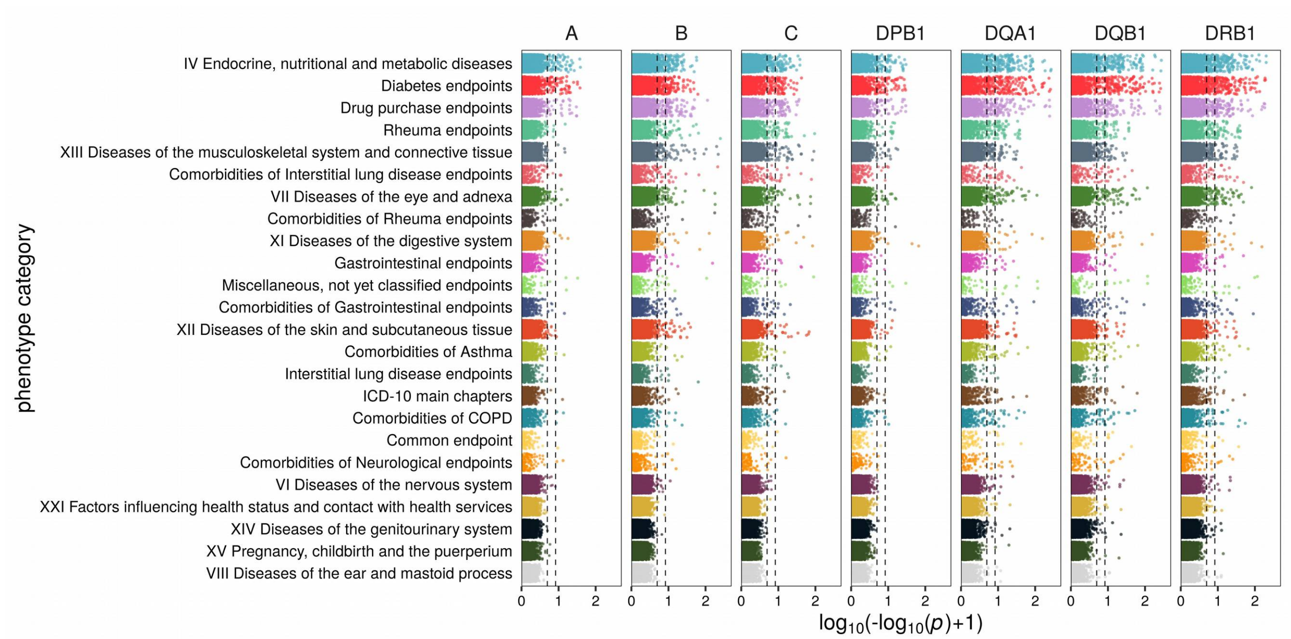


**Figure S1.** Overview of HLA allele associations in top phenotype categories. Distribution of association p-values for each analyzed HLA gene in the discovery cohort is shown on a double-logarithmic scale (x-axis). The top disease phenotype categories (y-axis) are shown in descending order of significant associations. The dashed vertical lines from left to right indicate FDR < 0.01 and genome-wide significance ($p < 5 \times 10^{-8}$) thresholds, correspondingly. The results reflect both the number of associated alleles and the number of phenotypes within the disease categories.

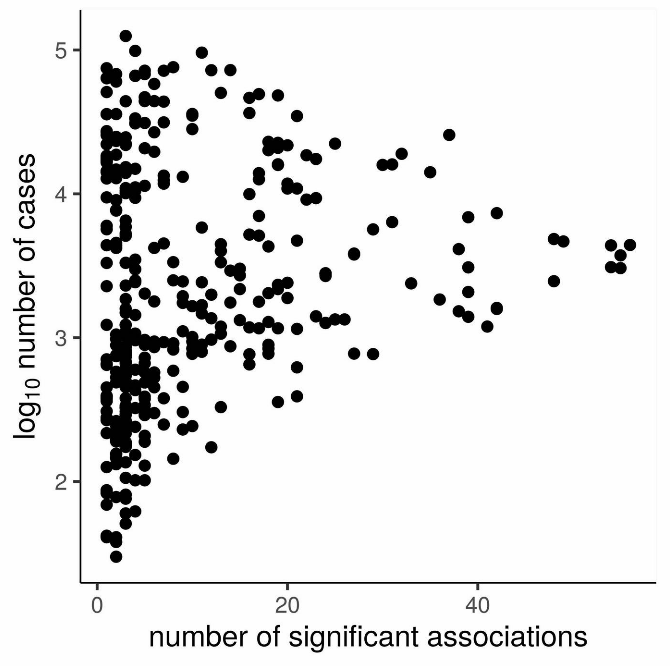


**Figure S2.** The number of cases in logarithmic scale vs. the number of observed significant HLA allele associations. Each data point represents a phenotype.

### Supplementary text

#### List of FinnGen members

FinnGen is funded by two grants from Business Finland (HUS 4685/31/2016 and UH 4386/31/2016) and twelve industry partners (AbbVie Inc, AstraZeneca UK Ltd, Biogen MA Inc, Celgene Corporation, Celgene International II Sàrl, Genentech Inc, GlaxoSmithKline, Janssen Biotech Inc. Maze Therapeutics Inc., Merck Sharp & Dohme Corp, Novartis, Pfizer Inc., Sanofi).

The Finnish biobanks are acknowledged for collecting the FinnGen samples: Auria Biobank (https://www.auria.fi/biopankki), THL Biobank (https://thl.fi/fi/web/thl-biopankki), Helsinki Biobank (https://www.terveyskyla.fi/helsinginbiopankki), Biobank Borealis of Northern Finland (https://www.oulu.fi/university/node/38474), Finnish Clinical Biobank Tampere (https://www.tays.fi/en-US/Research_and_development/Finnish_Clinical_Biobank_Tampere), Biobank of Eastern Finland (https://ita-suomenbiopankki.fi), Central Finland Biobank (https://www.ksshp.fi/fi-FI/Potilaalle/Biopankki), Finnish Red Cross Blood Service Biobank (https://www.veripalvelu.fi/verenluovutus/biopankkitoiminta) and Terveystalo Biobank (https://www.terveystalo.com/fi/Yritystietoa/Terveystalo-Biopankki/

Biopankki/). All Finnish Biobanks are members of BBMRI.fi infrastructure (www.bbmri.fi).

**Steering Committee**

| | |
|---|---|
| Aarno Palotie | Institute for Molecular Medicine Finland, HiLIFE, University of Helsinki, Finland |
| Mark Daly | Institute for Molecular Medicine Finland, HiLIFE, University of Helsinki, Finland |

**Pharmaceutical companies**

| | |
|---|---|
| Howard Jacob | Abbvie, Chicago, IL, United States |
| Athena Matakidou | Astra Zeneca, Cambridge, United Kingdom |
| Heiko Runz | Biogen, Cambridge, MA, United States |
| Sally John | Biogen, Cambridge, MA, United States |
| Robert Plenge | Celgene, Summit, NJ, United States |
| Mark McCarthy | Genentech, San Francisco, CA, United States |
| Julie Hunkapiller | Genentech, San Francisco, CA, United States |
| Meg Ehm | GlaxoSmithKline, Brentford, United Kingdom |
| Dawn Waterworth | GlaxoSmithKline, Brentford, United Kingdom |
| Caroline Fox | Merck, Kenilworth, NJ, United States |
| Anders Malarstig | Pfizer, New York, NY, United States |
| Kathy Klinger | Sanofi, Paris, France |
| Kathy Call | Sanofi, Paris, France |

**University of Helsinki & Biobanks**

| | |
|---|---|
| Tomi Mäkelä | HiLIFE, University of Helsinki, Finland, Finland |
| Jaakko Kaprio | Institute for Molecular Medicine Finland, HiLIFE, Helsinki, Finland, Finland |
| Petri Virolainen | Auria Biobank / Univ. of Turku / Hospital District of Southwest Finland, Turku, Finland |
| Kari Pulkki | Auria Biobank / Univ. of Turku / Hospital District of Southwest Finland, Turku, Finland |
| Terhi Kilpi | THL Biobank / Finnish Institute for Health and Welfare Helsinki, Finland |
| Markus Perola | THL Biobank / Finnish Institute for Health and Welfare Helsinki, Finland |

| | |
|---|---|
| Jukka Partanen | Finnish Red Cross Blood Service / Finnish Hematology Registry and Clinical Biobank, Helsinki, Finland |
| Anne Pitkäranta | Hospital District of Helsinki and Uusimaa, Helsinki, Finland |
| Riitta Kaarteenaho | Northern Finland Biobank Borealis / University of Oulu / Northern Ostrobothnia Hospital District, Oulu, Finland |
| Seppo Vainio | Northern Finland Biobank Borealis / University of Oulu / Northern Ostrobothnia Hospital District, Oulu, Finland |
| Kimmo Savinainen | Finnish Clinical Biobank Tampere / University of Tampere / Pirkanmaa Hospital District, Tampere, Finland |
| Veli-Matti Kosma | Biobank of Eastern Finland / University of Eastern Finland / Northern Savo Hospital District, Kuopio, Finland |
| Urho Kujala | Central Finland Biobank / University of Jyväskylä / Central Finland Health Care District, Jyväskylä, Finland |

**Other Experts/ Non-Voting Members**

| | |
|---|---|
| Outi Tuovila | Business Finland, Helsinki, Finland |
| Minna Hendolin | Business Finland, Helsinki, Finland |
| Raimo Pakkanen | Business Finland, Helsinki, Finland |

**Scientific Committee**

**Pharmaceutical companies**

| | |
|---|---|
| Jeff Waring | Abbvie, Chicago, IL, United States |
| Bridget Riley-Gillis | Abbvie, Chicago, IL, United States |
| Athena Matakidou | Astra Zeneca, Cambridge, United Kingdom |
| Heiko Runz | Biogen, Cambridge, MA, United States |
| Jimmy Liu | Biogen, Cambridge, MA, United States |
| Shameek Biswas | Celgene, Summit, NJ, United States |
| Julie Hunkapiller | Genentech, San Francisco, CA, United States |
| Dawn Waterworth | GlaxoSmithKline, Brentford, United Kingdom |
| Meg Ehm | GlaxoSmithKline, Brentford, United Kingdom |
| Dorothee Diogo | Merck, Kenilworth, NJ, United States |
| Caroline Fox | Merck, Kenilworth, NJ, United States |
| Anders Malarstig | Pfizer, New York, NY, United States |
| Catherine Marshall | Pfizer, New York, NY, United States |
| Xinli Hu | Pfizer, New York, NY, United States |
| Kathy Call | Sanofi, Paris, France |

| | |
|---|---|
| Kathy Klinger | Sanofi, Paris, France |
| Matthias Gossel | Sanofi, Paris, France |

**University of Helsinki & Biobanks**

| | |
|---|---|
| Samuli Ripatti | Institute for Molecular Medicine Finland, HiLIFE, University of Helsinki, Helsinki, Finland |
| Johanna Schleutker | Auria Biobank / Univ. of Turku / Hospital District of Southwest Finland, Turku, Finland |
| Markus Perola | THL Biobank / Finnish Institute for Health and Welfare Helsinki, Finland |
| Mikko Arvas | Finnish Red Cross Blood Service / Finnish Hematology Registry and Clinical Biobank, Helsinki, Finland |
| Olli Carpen | Hospital District of Helsinki and Uusimaa, Helsinki, Finland |
| Reetta Hinttala | Northern Finland Biobank Borealis / University of Oulu / Northern Ostrobothnia Hospital District, Oulu, Finland |
| Johannes Kettunen | Northern Finland Biobank Borealis / University of Oulu / Northern Ostrobothnia Hospital District, Oulu, Finland |
| Reijo Laaksonen | Finnish Clinical Biobank Tampere / University of Tampere / Pirkanmaa Hospital District, Tampere, Finland |
| Arto Mannermaa | Biobank of Eastern Finland / University of Eastern Finland / Northern Savo Hospital District, Kuopio, Finland |
| Juha Paloneva | Central Finland Biobank / University of Jyväskylä / Central Finland Health Care District, Jyväskylä, Finland |
| Urho Kujala | Central Finland Biobank / University of Jyväskylä / Central Finland Health Care District, Jyväskylä, Finland |

**Other Experts/ Non-Voting Members**

| | |
|---|---|
| Outi Tuovila | Business Finland, Helsinki, Finland |
| Minna Hendolin | Business Finland, Helsinki, Finland |
| Raimo Pakkanen | Business Finland, Helsinki, Finland |

**Clinical Groups**

**Neurology Group**

| | |
|---|---|
| Hilkka Soininen | Northern Savo Hospital District, Kuopio, Finland |
| Valtteri Julkunen | Northern Savo Hospital District, Kuopio, Finland |
| Anne Remes | Northern Ostrobothnia Hospital District, Oulu, Finland |
| Reetta Kälviäinen | Northern Savo Hospital District, Kuopio, Finland |
| Mikko Hiltunen | Northern Savo Hospital District, Kuopio, Finland |

| | |
|---|---|
| Jukka Peltola | Pirkanmaa Hospital District, Tampere, Finland |
| Pentti Tienari | Hospital District of Helsinki and Uusimaa, Helsinki, Finland |
| Juha Rinne | Hospital District of Southwest Finland, Turku, Finland |
| Adam Ziemann | Abbvie, Chicago, IL, United States |
| Jeffrey Waring | Abbvie, Chicago, IL, United States |
| Sahar Esmaeeli | Abbvie, Chicago, IL, United States |
| Nizar Smaoui | Abbvie, Chicago, IL, United States |
| Anne Lehtonen | Abbvie, Chicago, IL, United States |
| Susan Eaton | Biogen, Cambridge, MA, United States |
| Heiko Runz | Biogen, Cambridge, MA, United States |
| Sanni Lahdenperä | Biogen, Cambridge, MA, United States |
| Shameek Biswas | Celgene, Summit, NJ, United States |
| John Michon | Genentech, San Francisco, CA, United States |
| Geoff Kerchner | Genentech, San Francisco, CA, United States |
| Julie Hunkapiller | Genentech, San Francisco, CA, United States |
| Natalie Bowers | Genentech, San Francisco, CA, United States |
| Edmond Teng | Genentech, San Francisco, CA, United States |
| John Eicher | Merck, Kenilworth, NJ, United States |
| Vinay Mehta | Merck, Kenilworth, NJ, United States |
| Padhraig Gormley | Merck, Kenilworth, NJ, United States |
| Kari Linden | Pfizer, New York, NY, United States |
| Christopher Whelan | Pfizer, New York, NY, United States |
| Fanli Xu | GlaxoSmithKline, Brentford, United Kingdom |
| David Pulford | GlaxoSmithKline, Brentford, United Kingdom |

**Gastroenterology Group**

| | |
|---|---|
| Martti Färkkilä | Hospital District of Helsinki and Uusimaa, Helsinki, Finland |
| Sampsa Pikkarainen | Hospital District of Helsinki and Uusimaa, Helsinki, Finland |
| Airi Jussila | Pirkanmaa Hospital District, Tampere, Finland |
| Timo Blomster | Northern Ostrobothnia Hospital District, Oulu, Finland |
| Mikko Kiviniemi | Northern Savo Hospital District, Kuopio, Finland |
| Markku Voutilainen | Hospital District of Southwest Finland, Turku, Finland |
| Bob Georgantas | Abbvie, Chicago, IL, United States |

| | |
|---|---|
| Graham Heap | Abbvie, Chicago, IL, United States |
| Jeffrey Waring | Abbvie, Chicago, IL, United States |
| Nizar Smaoui | Abbvie, Chicago, IL, United States |
| Fedik Rahimov | Abbvie, Chicago, IL, United States |
| Anne Lehtonen | Abbvie, Chicago, IL, United States |
| Keith Usiskin | Celgene, Summit, NJ, United States |
| Joseph Maranville | Celgene, Summit, NJ, United States |
| Tim Lu | Genentech, San Francisco, CA, United States |
| Natalie Bowers | Genentech, San Francisco, CA, United States |
| Danny Oh | Genentech, San Francisco, CA, United States |
| John Michon | Genentech, San Francisco, CA, United States |
| Vinay Mehta | Merck, Kenilworth, NJ, United States |
| Kirsi Kalpala | Pfizer, New York, NY, United States |
| Melissa Miller | Pfizer, New York, NY, United States |
| Xinli Hu | Pfizer, New York, NY, United States |
| Linda McCarthy | GlaxoSmithKline, Brentford, United Kingdom |

**Rheumatology Group**

| | |
|---|---|
| Kari Eklund | Hospital District of Helsinki and Uusimaa, Helsinki, Finland |
| Antti Palomäki | Hospital District of Southwest Finland, Turku, Finland |
| Pia Isomäki | Pirkanmaa Hospital District, Tampere, Finland |
| Laura Pirilä | Hospital District of Southwest Finland, Turku, Finland |
| Oili Kaipiainen-Seppänen | Northern Savo Hospital District, Kuopio, Finland |
| Johanna Huhtakangas | Northern Ostrobothnia Hospital District, Oulu, Finland |
| Bob Georgantas | Abbvie, Chicago, IL, United States |
| Jeffrey Waring | Abbvie, Chicago, IL, United States |
| Fedik Rahimov | Abbvie, Chicago, IL, United States |
| Apinya Lertratanakul | Abbvie, Chicago, IL, United States |
| Nizar Smaoui | Abbvie, Chicago, IL, United States |
| Anne Lehtonen | Abbvie, Chicago, IL, United States |
| David Close | Astra Zeneca, Cambridge, United Kingdom |
| Marla Hochfeld | Celgene, Summit, NJ, United States |
| Natalie Bowers | Genentech, San Francisco, CA, United States |

| | |
|---|---|
| John Michon | Genentech, San Francisco, CA, United States |
| Dorothee Diogo | Merck, Kenilworth, NJ, United States |
| Vinay Mehta | Merck, Kenilworth, NJ, United States |
| Kirsi Kalpala | Pfizer, New York, NY, United States |
| Nan Bing | Pfizer, New York, NY, United States |
| Xinli Hu | Pfizer, New York, NY, United States |
| Jorge Esparza | Gordillo GlaxoSmithKline, Brentford, United Kingdom |
| Nina Mars | Institute for Molecular Medicine Finland, HiLIFE, University of Helsinki, Helsinki, Finland |

**Pulmonology Group**

| | |
|---|---|
| Tarja Laitinen | Pirkanmaa Hospital District, Tampere, Finland |
| Margit Pelkonen | Northern Savo Hospital District, Kuopio, Finland |
| Paula Kauppi | Hospital District of Helsinki and Uusimaa, Helsinki, Finland |
| Hannu Kankaanranta | Pirkanmaa Hospital District, Tampere, Finland |
| Terttu Harju | Northern Ostrobothnia Hospital District, Oulu, Finland |
| Nizar Smaoui | Abbvie, Chicago, IL, United States |
| David Close | Astra Zeneca, Cambridge, United Kingdom |
| Steven Greenberg | Celgene, Summit, NJ, United States |
| Hubert Chen | Genentech, San Francisco, CA, United States |
| Natalie Bowers | Genentech, San Francisco, CA, United States |
| John Michon | Genentech, San Francisco, CA, United States |
| Vinay Mehta | Merck, Kenilworth, NJ, United States |
| Jo Betts | GlaxoSmithKline, Brentford, United Kingdom |
| Soumitra Ghosh | GlaxoSmithKline, Brentford, United Kingdom |

**Cardiometabolic Diseases Group**

| | |
|---|---|
| Veikko Salomaa | Finnish Institute for Health and Welfare Helsinki, Finland |
| Teemu Niiranen | Finnish Institute for Health and Welfare Helsinki, Finland |
| Markus Juonala | Hospital District of Southwest Finland, Turku, Finland |
| Kaj Metsärinne | Hospital District of Southwest Finland, Turku, Finland |
| Mika Kähönen | Pirkanmaa Hospital District, Tampere, Finland |
| Juhani Junttila | Northern Ostrobothnia Hospital District, Oulu, Finland |
| Markku Laakso | Northern Savo Hospital District, Kuopio, Finland |

| | |
|---|---|
| Jussi Pihlajamäki | Northern Savo Hospital District, Kuopio, Finland |
| Juha Sinisalo | Hospital District of Helsinki and Uusimaa, Helsinki, Finland |
| Marja-Riitta Taskinen | Hospital District of Helsinki and Uusimaa, Helsinki, Finland |
| Tiinamaija Tuomi | Hospital District of Helsinki and Uusimaa, Helsinki, Finland |
| Jari Laukkanen | Central Finland Health Care District, Jyväskylä, Finland |
| Ben Challis | Astra Zeneca, Cambridge, United Kingdom |
| Andrew Peterson | Genentech, San Francisco, CA, United States |
| Julie Hunkapiller | Genentech, San Francisco, CA, United States |
| Natalie Bowers | Genentech, San Francisco, CA, United States |
| John Michon | Genentech, San Francisco, CA, United States |
| Dorothee Diogo | Merck, Kenilworth, NJ, United States |
| Audrey Chu | Merck, Kenilworth, NJ, United States |
| Vinay Mehta | Merck, Kenilworth, NJ, United States |
| Jaakko Parkkinen | Pfizer, New York, NY, United States |
| Melissa Miller | Pfizer, New York, NY, United States |
| Anthony Muslin | Sanofi, Paris, France |
| Dawn Waterworth | GlaxoSmithKline, Brentford, United Kingdom |

**Oncology Group**

| | |
|---|---|
| Heikki Joensuu | Hospital District of Helsinki and Uusimaa, Helsinki, Finland |
| Tuomo Meretoja | Hospital District of Helsinki and Uusimaa, Helsinki, Finland |
| Olli Carpen | Hospital District of Helsinki and Uusimaa, Helsinki, Finland |
| Lauri Aaltonen | Hospital District of Helsinki and Uusimaa, Helsinki, Finland |
| Annika Auranen | Pirkanmaa Hospital District, Tampere, Finland |
| Peeter Karihtala | Northern Ostrobothnia Hospital District, Oulu, Finland |
| Saila Kauppila | Northern Ostrobothnia Hospital District, Oulu, Finland |
| Päivi Auvinen | Northern Savo Hospital District, Kuopio, Finland |
| Klaus Elenius | Hospital District of Southwest Finland, Turku, Finland |
| Relja Popovic | Abbvie, Chicago, IL, United States |
| Jeffrey Waring | Abbvie, Chicago, IL, United States |
| Bridget Riley-Gillis | Abbvie, Chicago, IL, United States |
| Anne Lehtonen | Abbvie, Chicago, IL, United States |
| Athena Matakidou | Astra Zeneca, Cambridge, United Kingdom |

| | |
|---|---|
| Jennifer Schutzman | Genentech, San Francisco, CA, United States |
| Julie Hunkapiller | Genentech, San Francisco, CA, United States |
| Natalie Bowers | Genentech, San Francisco, CA, United States |
| John Michon | Genentech, San Francisco, CA, United States |
| Vinay Mehta | Merck, Kenilworth, NJ, United States |
| Andrey Loboda | Merck, Kenilworth, NJ, United States |
| Aparna Chhibber | Merck, Kenilworth, NJ, United States |
| Heli Lehtonen | Pfizer, New York, NY, United States |
| Stefan McDonough | Pfizer, New York, NY, United States |
| Marika Crohns | Sanofi, Paris, France |
| Diptee Kulkarni | GlaxoSmithKline, Brentford, United Kingdom |

**Opthalmology Group**

| | |
|---|---|
| Kai Kaarniranta | Northern Savo Hospital District, Kuopio, Finland |
| Joni Turunen | Hospital District of Helsinki and Uusimaa, Helsinki, Finland |
| Terhi Ollila | Hospital District of Helsinki and Uusimaa, Helsinki, Finland |
| Sanna Seitsonen | Hospital District of Helsinki and Uusimaa, Helsinki, Finland |
| Hannu Uusitalo | Pirkanmaa Hospital District, Tampere, Finland |
| Vesa Aaltonen | Hospital District of Southwest Finland, Turku, Finland |
| Hannele Uusitalo-Järvinen | Pirkanmaa Hospital District, Tampere, Finland |
| Marja Luodonpää | Northern Ostrobothnia Hospital District, Oulu, Finland |
| Nina Hautala | Northern Ostrobothnia Hospital District, Oulu, Finland |
| Heiko Runz | Biogen, Cambridge, MA, United States |
| Erich Strauss | Genentech, San Francisco, CA, United States |
| Natalie Bowers | Genentech, San Francisco, CA, United States |
| Hao Chen | Genentech, San Francisco, CA, United States |
| John Michon | Genentech, San Francisco, CA, United States |
| Anna Podgornaia | Merck, Kenilworth, NJ, United States |
| Vinay Mehta | Merck, Kenilworth, NJ, United States |
| Dorothee Diogo | Merck, Kenilworth, NJ, United States |
| Joshua Hoffman | GlaxoSmithKline, Brentford, United Kingdom |

**Dermatology Group**

| | |
|---|---|
| Kaisa Tasanen | Northern Ostrobothnia Hospital District, Oulu, Finland |
| Laura Huilaja | Northern Ostrobothnia Hospital District, Oulu, Finland |
| Katariina Hannula-Jouppi | Hospital District of Helsinki and Uusimaa, Helsinki, Finland |
| Teea Salmi Pirkanmaa | Hospital District, Tampere, Finland |
| Sirkku Peltonen | Hospital District of Southwest Finland, Turku, Finland |
| Leena Koulu | Hospital District of Southwest Finland, Turku, Finland |
| Ilkka Harvima | Northern Savo Hospital District, Kuopio, Finland |
| Kirsi Kalpala | Pfizer, New York, NY, United States |
| Ying Wu | Pfizer, New York, NY, United States |
| David Choy | Genentech, San Francisco, CA, United States |
| John Michon | Genentech, San Francisco, CA, United States |
| Nizar Smaoui | Abbvie, Chicago, IL, United States |
| Fedik Rahimov | Abbvie, Chicago, IL, United States |
| Anne Lehtonen | Abbvie, Chicago, IL, United States |
| Dawn Waterworth | GlaxoSmithKline, Brentford, United Kingdom |

**FinnGen Teams**

**Administration Team**

| | |
|---|---|
| Anu Jalanko | Institute for Molecular Medicine Finland, HiLIFE, University of Helsinki, Finland |
| Risto Kajanne | Institute for Molecular Medicine Finland, HiLIFE, University of Helsinki, Finland |
| Mervi Aavikko | Institute for Molecular Medicine Finland, HiLIFE, University of Helsinki, Finland |
| Manuel González Jiménez | Institute for Molecular Medicine Finland, HiLIFE, University of Helsinki, Finland |

**Communication**

| | |
|---|---|
| Mari Kaunisto | Institute for Molecular Medicine Finland, HiLIFE, University of Helsinki, Finland |

**Analysis Team**

| | |
|---|---|
| Justin Wade Davis | Abbvie, Chicago, IL, United States |
| Bridget Riley-Gillis | Abbvie, Chicago, IL, United States |
| Danjuma Quarless | Abbvie, Chicago, IL, United States |

| | |
|---|---|
| Slavé Petrovski | Astra Zeneca, Cambridge, United Kingdom |
| Jimmy Liu | Biogen, Cambridge, MA, United States |
| Chia-Yen Chen | Biogen, Cambridge, MA, United States |
| Paola Bronson | Biogen, Cambridge, MA, United States |
| Robert Yang | Celgene, Summit, NJ, United States |
| Joseph Maranville | Celgene, Summit, NJ, United States |
| Shameek Biswas | Celgene, Summit, NJ, United States |
| Diana Chang | Genentech, San Francisco, CA, United States |
| Julie Hunkapiller | Genentech, San Francisco, CA, United States |
| Tushar Bhangale | Genentech, San Francisco, CA, United States |
| Natalie Bowers | Genentech, San Francisco, CA, United States |
| Dorothee Diogo | Merck, Kenilworth, NJ, United States |
| Emily Holzinger | Merck, Kenilworth, NJ, United States |
| Padhraig Gormley | Merck, Kenilworth, NJ, United States |
| Xulong Wang | Merck, Kenilworth, NJ, United States |
| Xing Chen | Pfizer, New York, NY, United States |
| Åsa Hedman | Pfizer, New York, NY, United States |
| Kirsi Auro | GlaxoSmithKline, Brentford, United Kingdom |
| Clarence Wang | Sanofi, Paris, France |
| Ethan Xu | Sanofi, Paris, France |
| Franck Auge | Sanofi, Paris, France |
| Clement Chatelain | Sanofi, Paris, France |
| Mitja Kurki | Institute for Molecular Medicine Finland, HiLIFE, University of Helsinki, Finland / Broad Institute, Cambridge, MA, United States |
| Samuli Ripatti | Institute for Molecular Medicine Finland, HiLIFE, University of Helsinki, Finland |
| Mark Daly | Institute for Molecular Medicine Finland, HiLIFE, University of Helsinki, Finland |
| Juha Karjalainen | Institute for Molecular Medicine Finland, HiLIFE, University of Helsinki, Finland / Broad Institute, Cambridge, MA, United States |
| Aki Havulinna | Institute for Molecular Medicine Finland, HiLIFE, University of Helsinki, Finland |

| | |
|---|---|
| Anu Jalanko | Institute for Molecular Medicine Finland, HiLIFE, University of Helsinki, Finland |
| Kimmo Palin | University of Helsinki, Helsinki, Finland |
| Priit Palta | Institute for Molecular Medicine Finland, HiLIFE, University of Helsinki, Finland |
| Pietro Della Briotta Parolo | Institute for Molecular Medicine Finland, HiLIFE, University of Helsinki, Finland |
| Wei Zhou | Broad Institute, Cambridge, MA, United States |
| Susanna Lemmelä | Institute for Molecular Medicine Finland, HiLIFE, University of Helsinki, Finland |
| Manuel Rivas | University of Stanford, Stanford, CA, United States |
| Jarmo Harju | Institute for Molecular Medicine Finland, HiLIFE, University of Helsinki, Finland |
| Aarno Palotie | Institute for Molecular Medicine Finland, HiLIFE, University of Helsinki, Finland |
| Arto Lehisto | Institute for Molecular Medicine Finland, HiLIFE, University of Helsinki, Finland |
| Andrea Ganna | Institute for Molecular Medicine Finland, HiLIFE, University of Helsinki, Finland |
| Vincent Llorens | Institute for Molecular Medicine Finland, HiLIFE, University of Helsinki, Finland |
| Antti Karlsson | Auria Biobank / Univ. of Turku / Hospital District of Southwest Finland, Turku, Finland |
| Kati Kristiansson | THL Biobank / Finnish Institute for Health and Welfare Helsinki, Finland |
| Mikko Arvas | Finnish Red Cross Blood Service / Finnish Hematology Registry and Clinical Biobank, Helsinki, Finland |
| Kati Hyvärinen | Finnish Red Cross Blood Service / Finnish Hematology Registry and Clinical Biobank, Helsinki, Finland |
| Jarmo Ritari | Finnish Red Cross Blood Service / Finnish Hematology Registry and Clinical Biobank, Helsinki, Finland |
| Tiina Wahlfors | Finnish Red Cross Blood Service / Finnish Hematology Registry and Clinical Biobank, Helsinki, Finland |
| Miika Koskinen | Hospital District of Helsinki and Uusimaa, Helsinki, Finland BB/HUS/Univ Hosp Districts |
| Olli Carpen | Hospital District of Helsinki and Uusimaa, Helsinki, Finland BB/HUS/Univ Hosp Districts |

| | |
|---|---|
| Johannes Kettunen | Northern Finland Biobank Borealis / University of Oulu / Northern Ostrobothnia Hospital District, Oulu, Finland |
| Katri Pylkäs | Northern Finland Biobank Borealis / University of Oulu / Northern Ostrobothnia Hospital District, Oulu, Finland |
| Marita Kalaoja | Northern Finland Biobank Borealis / University of Oulu / Northern Ostrobothnia Hospital District, Oulu, Finland |
| Minna Karjalainen | Northern Finland Biobank Borealis / University of Oulu / Northern Ostrobothnia Hospital District, Oulu, Finland |
| Tuomo Mantere | Northern Finland Biobank Borealis / University of Oulu / Northern Ostrobothnia Hospital District, Oulu, Finland |
| Eeva Kangasniemi | Finnish Clinical Biobank Tampere / University of Tampere / Pirkanmaa Hospital District, Tampere, Finland |
| Sami Heikkinen | Biobank of Eastern Finland / University of Eastern Finland / Northern Savo Hospital District, Kuopio, Finland |
| Arto Mannermaa | Biobank of Eastern Finland / University of Eastern Finland / Northern Savo Hospital District, Kuopio, Finland |
| Eija Laakkonen | Central Finland Biobank / University of Jyväskylä / Central Finland Health Care District, Jyväskylä, Finland |
| Juha Kononen | Central Finland Biobank / University of Jyväskylä / Central Finland Health Care District, Jyväskylä, Finland |

**Sample Collection Coordination**

| | |
|---|---|
| Anu Loukola | Hospital District of Helsinki and Uusimaa, Helsinki, Finland |

**Sample Logistics**

| | |
|---|---|
| Päivi Laiho | THL Biobank / Finnish Institute for Health and Welfare Helsinki, Finland |
| Tuuli Sistonen | THL Biobank / Finnish Institute for Health and Welfare Helsinki, Finland |
| Essi Kaiharju | THL Biobank / Finnish Institute for Health and Welfare Helsinki, Finland |
| Markku Laukkanen | THL Biobank / Finnish Institute for Health and Welfare Helsinki, Finland |
| Elina Järvensivu | THL Biobank / Finnish Institute for Health and Welfare Helsinki, Finland |
| Sini Lähteenmäki | THL Biobank / Finnish Institute for Health and Welfare Helsinki, Finland |

| | |
|---|---|
| Lotta Männikkö | THL Biobank / Finnish Institute for Health and Welfare Helsinki, Finland |
| Regis Wong | THL Biobank / Finnish Institute for Health and Welfare Helsinki, Finland |

**Registry Data Operations**

| | |
|---|---|
| Kati Kristiansson | THL Biobank / Finnish Institute for Health and Welfare Helsinki, Finland |
| Hannele Mattsson | THL Biobank / Finnish Institute for Health and Welfare Helsinki, Finland |
| Susanna Lemmelä | Institute for Molecular Medicine Finland, HiLIFE, University of Helsinki, Finland |
| Tero Hiekkalinna | THL Biobank / Finnish Institute for Health and Welfare Helsinki, Finland |
| Manuel González Jiménez | THL Biobank / Finnish Institute for Health and Welfare Helsinki, Finland |

**Genotyping**

| | |
|---|---|
| Kati Donner | Institute for Molecular Medicine Finland, HiLIFE, University of Helsinki, Finland |

**Sequencing Informatics**

| | |
|---|---|
| Priit Palta | Institute for Molecular Medicine Finland, HiLIFE, University of Helsinki, Finland |
| Kalle Pärn | Institute for Molecular Medicine Finland, HiLIFE, University of Helsinki, Finland |
| Javier Nunez-Fontarnau | Institute for Molecular Medicine Finland, HiLIFE, University of Helsinki, Finland |

**Data Management and IT Infrastructure**

| | |
|---|---|
| Jarmo Harju | Institute for Molecular Medicine Finland, HiLIFE, University of Helsinki, Finland |
| Elina Kilpeläinen | Institute for Molecular Medicine Finland, HiLIFE, University of Helsinki, Finland |
| Timo P. Sipilä | Institute for Molecular Medicine Finland, HiLIFE, University of Helsinki, Finland |
| Georg Brein | Institute for Molecular Medicine Finland, HiLIFE, University of Helsinki, Finland |
| Alexander Dada | Institute for Molecular Medicine Finland, HiLIFE, University of Helsinki, Finland |

| | |
|---|---|
| Ghazal Awaisa | Institute for Molecular Medicine Finland, HiLIFE, University of Helsinki, Finland |
| Anastasia Shcherban | Institute for Molecular Medicine Finland, HiLIFE, University of Helsinki, Finland |
| Tuomas Sipilä | Institute for Molecular Medicine Finland, HiLIFE, University of Helsinki, Finland |

**Clinical Endpoint Development**

| | |
|---|---|
| Hannele Laivuori | Institute for Molecular Medicine Finland, HiLIFE, University of Helsinki, Finland |
| Aki Havulinna | Institute for Molecular Medicine Finland, HiLIFE, University of Helsinki, Finland |
| Susanna Lemmelä | Institute for Molecular Medicine Finland, HiLIFE, University of Helsinki, Finland |
| Tuomo Kiiskinen | Institute for Molecular Medicine Finland, HiLIFE, University of Helsinki, Finland |

**Trajectory Team**

| | |
|---|---|
| Tarja Laitinen | Pirkanmaa Hospital District, Tampere, Finland |
| Harri Siirtola | University of Tampere, Tampere, Finland |
| Javier Gracia Tabuenca | University of Tampere, Tampere, Finland |

**Biobank Directors**

| | |
|---|---|
| Lila Kallio | Auria Biobank, Turku, Finland |
| Sirpa Soini | THL Biobank, Helsinki, Finland |
| Jukka Partanen | Blood Service Biobank, Helsinki, Finland |
| Kimmo Pitkänen | Helsinki Biobank, Helsinki, Finland |
| Seppo Vainio | Northern Finland Biobank Borealis, Oulu, Finland |
| Kimmo Savinainen | Tampere Biobank, Tampere, Finland |
| Veli-Matti Kosma | Biobank of Eastern Finland, Kuopio, Finland |
| Teijo Kuopio | Central Finland Biobank, Jyväskylä, Finland |
